# Growth charts for pontocerebellar hypoplasia type 2A

**DOI:** 10.1101/2024.06.23.24307757

**Authors:** Alice Kuhn, Maren Hackenberg, Anna-Lena Klauser, Antonia Herrmann, Julia Matilainen, Simone Mayer, Saskia Frölich, Ingeborg Krägeloh-Mann, Samuel Groeschel, Wibke G. Janzarik

## Abstract

**Introduction:** Pontocerebellar hypoplasia type 2A (PCH2A) is a rare, autosomal recessive disease, caused by a homozygous pathogenic variant in the gene *TSEN54* (c.919G>A, p.A307S). Apart from the characteristic pontocerebellar hypoplasia in MRI, PCH2A is clinically characterized by a dyskinetic movement disorder, severe neurodevelopment delay, progressive microcephaly, and, less well recognized, failure to thrive. Additional symptoms such as seizures, gastrointestinal or respiratory problems are common. The aim of this study was to document growth data of PCH2A patients, calculate growth charts for height, weight, body mass index (BMI) and head circumference (hc), and compare these to German reference charts.

**Patients and methods:** In total, data of 65 patients with genetically confirmed PCH2A were included in the study. Growth data were collected retrospectively from medical reports and a parent questionnaire. Disease-specific growth charts were prepared using gamlss package in R. Sex-disaggregated growth charts for PCH2A were compared to German reference data from the KiGGs study.

**Results:** Height and weight of patients with PCH2A were within the normal range at birth. Mean weight was significantly lower from the age of 3 months onwards, and mean height at the age of 6 months in patients with PCH2A, both, females and males. Mean BMI was statistically lower in patients at the age 4 months. Compared to reference values, mean head circumference of patients with PCH2A was significantly below average at birth, and all patients showed severe and progressive microcephaly in the further course.

**Conclusion:** In line with previous reports, patients with PCH2A typically exhibit progressive microcephaly, and frequently fail to thrive during infancy. Disease-specific growth charts of pediatric patients with PCH2A are provided as a helpful tool to monitor height, weight, BMI and head circumference of affected children.

## Introduction

The term Pontocerebellar hypoplasia (PCH) was initially established for congenital neurodegenerative diseases with pontocerebellar hypoplasia as characteristic anatomical feature (1). MRI of patients with Pontocerebellar hypoplasia type 2A (PCH2A) show prominent pontocerebellar hypoplasia with a “dragonfly” appearance, and progressive neocortical atrophy (2, 3). A considerable number of different types of PCH has been described with PCH2A, although still rare, being the most common type of PCH with less than 1 in 200 000 births (4, 5).

PCH2A refers to a homogeneous group of patients, genetically defined by a specific homozygous pathogenic variant of the gene *TSEN54* (c.919G>T; p.A307S), most likely due to a founder effect, and is inherited in an autosomal-recessive manner (4). TSEN54 is a subunit of the TSEN complex, which functions as a tRNA splicing endonuclease for intron-containing pre-tRNAs, and participates in pre-mRNA-3‘end formation (6). The TSEN complex is likely to play an important role during neuronal development, especially in the infratentorial region, as pathogenic variants result in congenital pontocerebellar hypoplasia and severe neurological symptoms (4).

Patients with PCH2A typically show profound neurodevelopmental impairment and a characteristic movement disorder with dyskinesia and/or chorea, often accompanied by dystonic attacks and/or spasticity (5). In many affected patients, abnormal movements are already observed during the neonatal period, as well as feeding difficulties (5). During the further course of the disease, epileptic seizures often occur, which can be difficult to treat (5). Progressive microcephaly has been reported as a major clinical characteristic of PCH2A, but also height and weight gain are often reduced (5). Due to dysphagia, patients often require percutaneous endoscopic gastrostomy (PEG) for feeding (5).

Feeding problems are a common finding in severe neurodevelopmental disorders (7). Disease-specific growth charts help to monitor the growth of individual patients in comparison with other affected patients of the same age group (8). For several genetic syndromes, such as Down syndrome, Turner syndrome, Achondroplasia or Mucopolysaccharidosis type III (MPS III), disease-specific growth charts have already been published (9–13). In a previous natural history study of patients with PCH2A, failure to thrive has been observed in addition to progressive microcephaly (5). However, to date, no PCH2A-specific growth charts are available. The aim of this study was to provide growth charts for height, weight, body mass index (BMI) and head circumference (hc) for children with PCH2A (0-18 years of age) living predominantly in Germany, and compare those to the German KiGGS (Kinder-und Jugendgesundheits-Survey) reference percentiles for anthropometric measurements (14, 15).

## Methods

### Patients

Data were collected retrospectively from a cohort of patients with genetically proven PCH2A, which were recruited via the German PCH patient network (PCH-Familie e.V.), and included data from the first natural history study (5). We retrospectively collected all available data from medical records and a parent questionnaire regarding height, weight, and hc, and subsequently calculated BMI. For pre-term born children, birth measurements were excluded from the analysis, and corrected age was used for all further measurements. Written informed consent was given by the parents in all cases. The study was approved by the ethics committee of the University of Freiburg in November 2020 (no. 20-1040); the first natural history study was approved by the ethics committee of the University of Tübingen (no. 105/2012BO2).

### Statistical analysis

Growth charts for PCH2A were plotted from birth to age 18 years for all body measurements. Mean height, weight, BMI, and hc were compared in a sex-disaggregated manner to the mean measurements of children from a healthy German reference population from the KiGGs survey (14, 15). Descriptive data was calculated using IBM SPSS Statistics and R core team (16, 17). For comparison, a one sample t-test or Wilcoxon test were used, depending on whether the assumption of normal distribution was fulfilled or not. To calculate difference in survival between male and female participants, Kaplan-Meier estimate and log-rank test were calculated. A p value < 0.05 was considered statistically significant.

To calculate the growth charts, LMS models, i.e. lambda, mu, sigma, were used (18). The LMS method allows to fit smooth centile curves to given data. It is based on the assumption that the distribution of measurements changing according to age can be modelled by suitable transformation of the median, coefficient of variation and skewness of the distribution (19). Specifically, the LMS method implement within the gamlss package (Generalised Additive Models for Location, Scale and Shape), version 5.4-10, in R core team was applied (16, 20, 21). The LMS method within the gamlss package is based on Box-Cox transformations, where the lambda (λ) corresponds to the skewness and is expressed as power in the Box-Cox transformation, mu (μ) corresponds to the Median and sigma (σ) to the generalized coefficient of variation (18–21). For each body measurement, three slightly different models, all modifications of the Box-Cox transformation, were fitted to the data. The available models are called Box-Cox Cole and Green distribution (BCCGo), Box-Cox Power Exponential distribution (BCPEo) and Box-Cox t distribution (BCTo) (20, 21). All default-parameters of the models, except “trans.x” and “k” were accepted. “Trans.x” describes the power transformation along the x-axis. Generally, body measurements show a skewed distribution over time. Setting “trans.x” as “TRUE” instead of “FALSE” (the default) was used to apply a transformation to the x-axis to normalise this skewed distribution (22). “K” describes a penalty in the calculation of the generalised Akaike information criterion, which is applied internally in the LMS method to evaluate goodness of fit and model convergence. (20, 21). The parameter was set to 3 instead of 2 (the default) according to the recommendation of Stasinopoulos et al., which is based on an automatic hyperparameter optimisation proposed in Rigby and Stasinopoulos (20, 23). To consider all available data points, a negligible random small value was added as jittering to every age value (24, 25). Growth charts were reviewed for medical plausibility to evaluate the model fit, and worm plots and q-statistics were performed (26). For q-statistics, recommended intervals were accepted, while for worm plots, interval-range was set to 9.

We applied the following criteria for fitting the gamlss models. Only models reaching convergence were accepted. If convergence could not be reached in BCCGo, BPEo, and BCTo, “k” was set back to the default (k=2). In some cases, even with “k” set to 2, no model convergence was achieved. In these instances, a normal model was used, which fits a normal distribution to the data, considering only mu and sigma. Consequently, it corresponds to a linear regression model (27). Regarding height, weight, and BMI, the BCTo model exhibited the best fit for data not specific to gender. Hc was calculated using the BCCGo model. Sex-disaggregated growth charts for height were calculated using the BCCGo model for females and the normal model for males. Concerning weight, the normal model was used to calculate female growth charts, and the BCCGo model was used for male growth charts. For the calculation of BMI, BCCGo was fitted to female data, whereas the BCTo model was fitted to male data. Sex-disaggregated growth charts for hc were calculated using the BCTo model with “k” set to 2 for female data and the BCCGo model for male data. Generally, we observed that the fitted centiles using various models within the GAMLSS family produced very similar results, indicating robustness of our approach and results.

## Results

### Study population

In total, data of 65 patients with genetically proven PCH2A were included (table 1). Of these, 33 patients had participated in the first natural history study (5). We received updated information of 21 of the 33 patients about growth and survival, and included data regarding survival of 5 additional patients of the first natural history study. No follow-up information was available in 7 patients due to loss to follow-up. Of the 65 patients, 56 originated from Germany, and nine additional patients came from Switzerland, Austria, and Bulgaria. Among the patients, there were 9 pairs of affected siblings. Gender was equally distributed. Three patients (all male) were born pre-term (gestational age 36+6, 36+0, and 30+4 weeks, respectively). For them, birth measurements were excluded from the analysis, and corrected age was used for all further measurements. Seven out of 33 female patients (21.2 %), and seven out of 32 male patients (21.9 %), respectively reached survival up to 15 years. No significant difference in survival was found between female and male patients with PCH2A (p=0.4).

**table 1:**
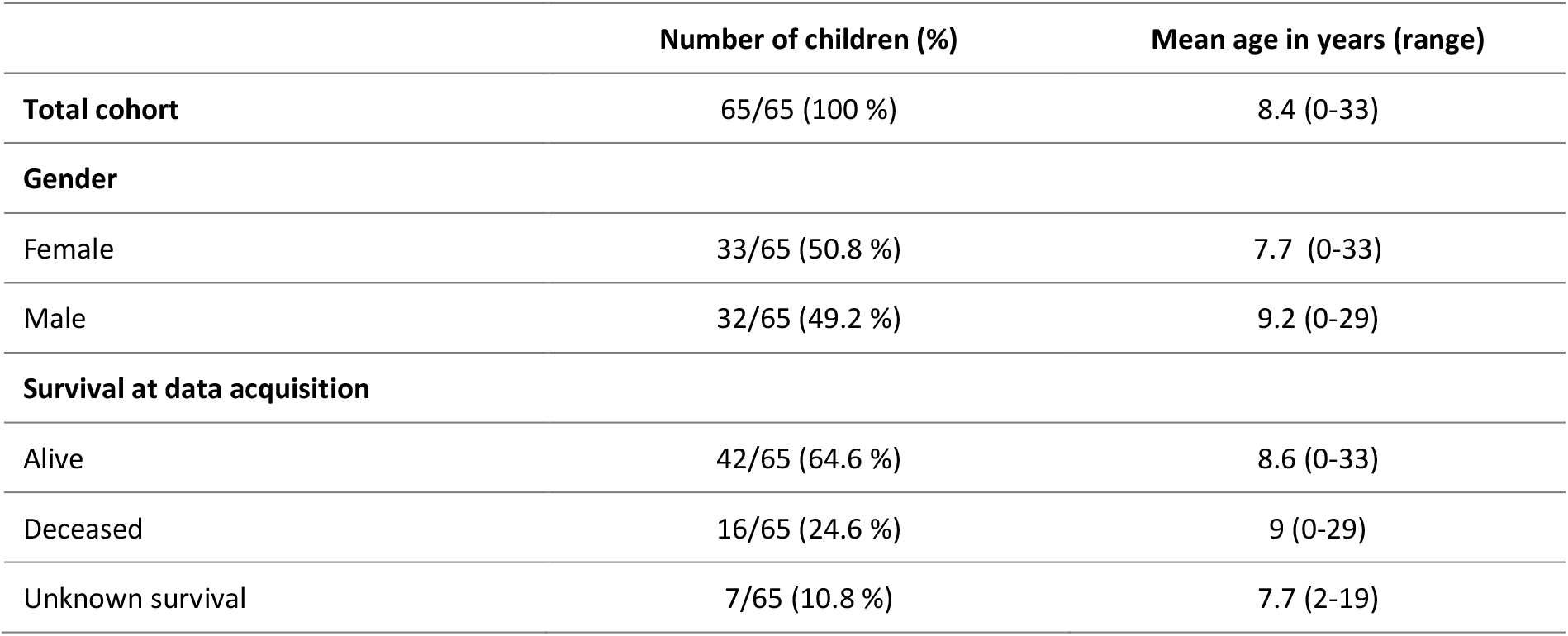
Study population.

|  | Number of children (%) | Mean age in years (range) |
| --- | --- | --- |
| **Total cohort** | 65/65 (100 %) | 8.4 (0-33) |
| **Gender** |  |  |
| Female | 33/65 (50.8 %) | 7.7 (0-33) |
| Male | 32/65 (49.2 %) | 9.2 (0-29) |
| **Survival at data acquisition** |  |  |
| Alive | 42/65 (64.6 %) | 8.6 (0-33) |
| Deceased | 16/65 (24.6 %) | 9 (0-29) |
| Unknown survival | 7/65 (10.8 %) | 7.7 (2-19) |

### Height

Overall, 643 individual measurements for height were collected from age 0-18 years, i.e. 353 measurements from female patients and 290 from male patients. The mean number of available measurements per patient was 9.8 (1-22). For the age span 0-2 years, 426 measurements were available, from 2-18 years of age, additional 217 measurements were available. Between 15-18 years of age, 11 measurements of female patients, and 6 measurements of male patients were included. Due to the more extensive data within the first two years of life, graphical and statistical comparison to the reference group was restricted to this age group.

For patients with PCH2A 0-18 years of age, height percentiles were calculated without sex-disaggregation (figure 1). However, individual measurements of female and male patients, marked in different colours, were included in the growth chart for comparison. For patients with PCH2A 0-2 years of age, sex-disaggregated percentiles are displayed in comparison to the reference group of healthy children (figure 2). At birth, mean height was 51.4 cm (SD: 2.16 cm) in female patients, and 52 cm (SD: 2.34 cm) in male patients, respectively. Both values did not differ significantly from the healthy reference group at birth (female: Z=-0.14, p=0.89; male: Z=0, p>0.99). With increasing age, height measurements showed a progressive deviation from the reference values, which reached statistical significance at 4 months of age in female (Z=-2.66, p=0.021), and at 6 months in male patients with PCH2A (t(17)=-2.16, p=0.046). In female patients, height remained below the healthy reference population with statistical significance from 6 months onwards (t(18)=-3.56, p=0.0022), and in male patients from 12 months onward (t(11)=-3.57, p=0.0044).

**figure 1:**
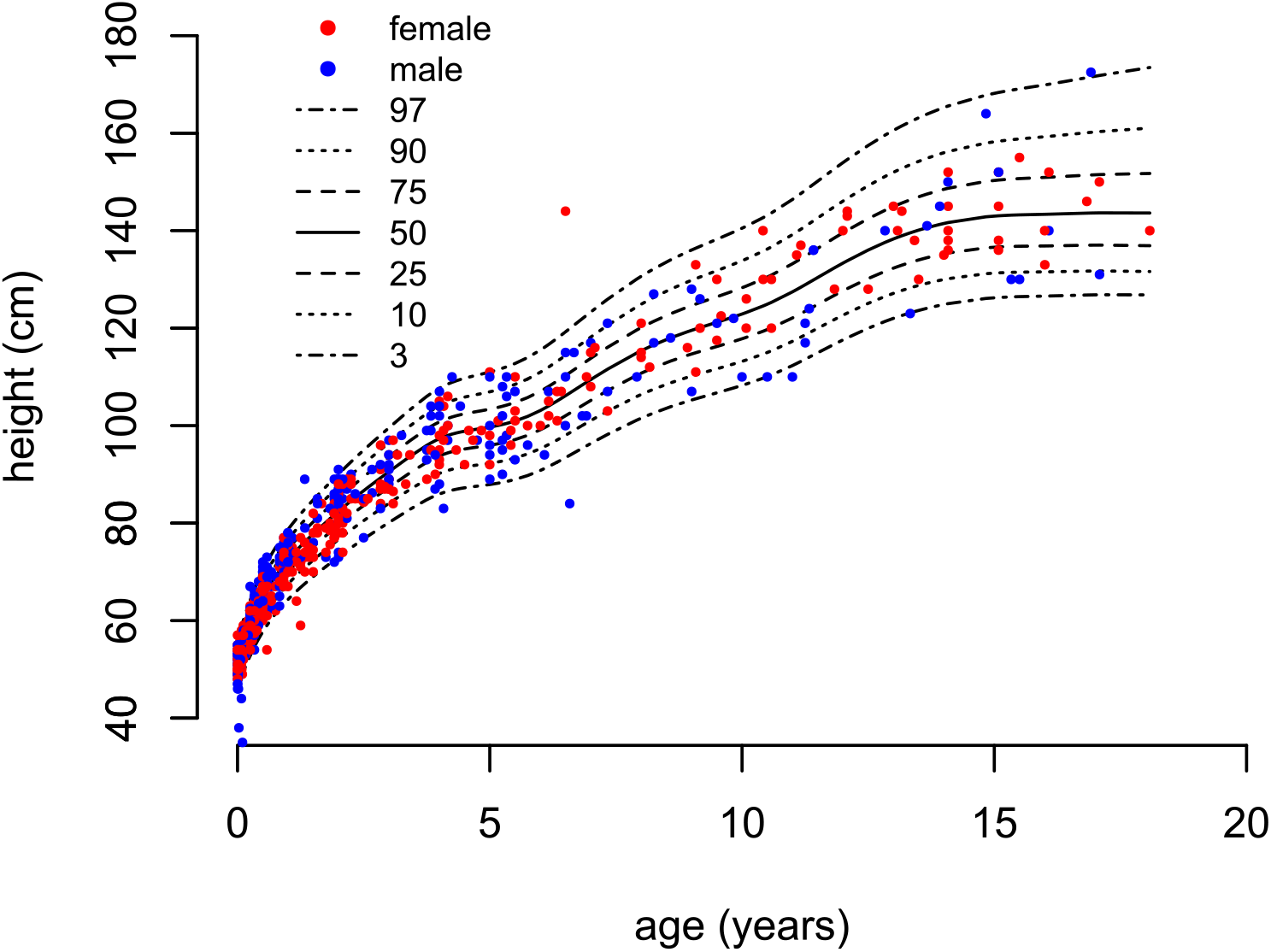
PCH2A-specific growth charts (0-18 years). BCTo model.

**figure 2:**
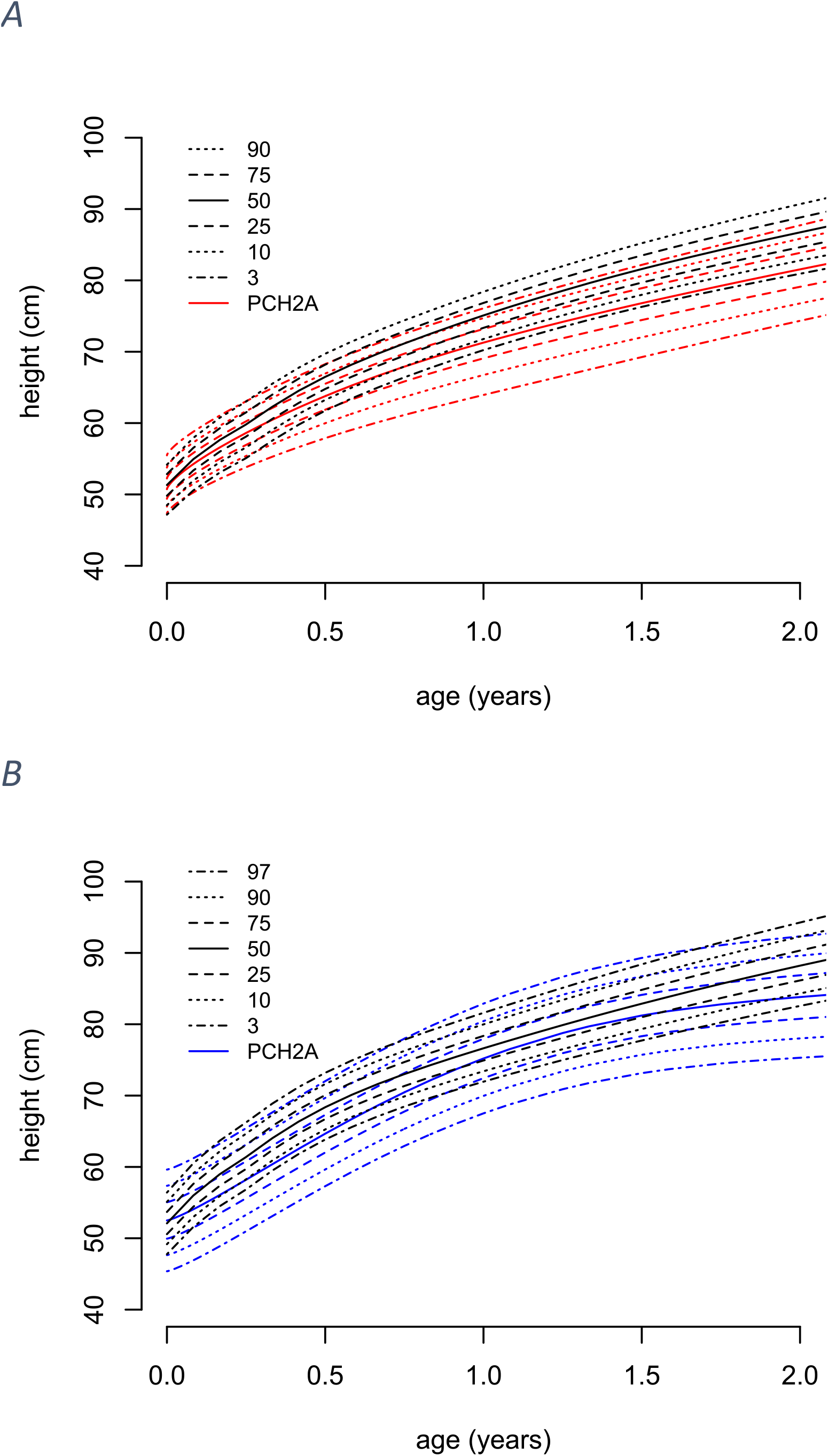
Sex-disaggregated growth charts for patients with PCH2A versus a healthy reference population (black, KiGGs) from 0-2 years; A females (red, 233 measurements, BCCGo nodel); B males (blue, 193 measurements, Normal model)

### Weight

Overall, 708 individual measurements for weight were collected from age 0-18 years, i.e. 385 measurements from female patients and 323 from male patients. The mean number of available measurements per patient was 10.9 (1-23). From 0-2 years, 452 measurements were available, and from 2-18 years of age, additional 256 measurements were available. In the age group 15-18 years, 12 measurements of female patients, and 9 measurements of male patients were included. Due to the more extensive data within the first 2 years of life, graphical and statistical comparison to the reference group was only performed for this age group.

For patients with PCH2A 0-18 years of age, weight percentiles were calculated without sex-disaggregation (figure 3). For comparison, the weight chart includes differently coloured individual measurements of female and male patients, respectively. For patients with PCH2A age 0-2 years, sex-disaggregated percentiles are displayed in comparison to the reference group of healthy children (figure 4).At birth, mean weight was 3390 g (SD: 430 g) in female patients, and 3530 g (SD: 403 g) in male patients. Both values did not differ significantly from the healthy reference group at birth (female: t(32)=0.02: p=0.98; male: t(28)=0.035; p=0.97). Weight measurements progressively deviated from the reference values over time. The difference to the reference values of healthy children reached statistical significance at the age of 3 months in female patients (t(13)=-3.44, p=0.0044), and at the age of 1 month in male patients (t(11)=-2.36, p=0.038), and remained statistically significant from then on.

**figure 3:**
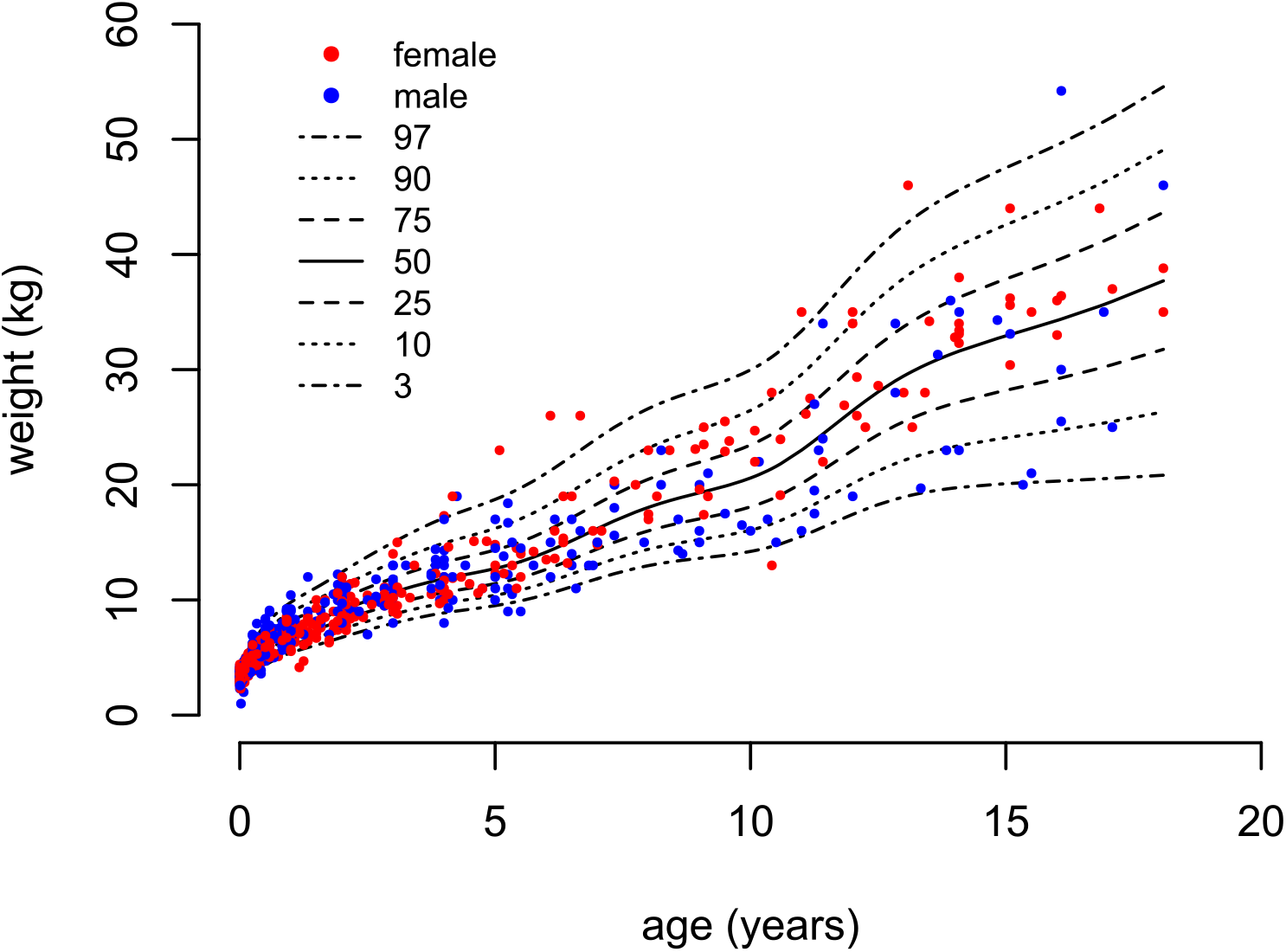
PCH2A-specific weight charts (0-18 years). BCTo model.

**figure 4:**
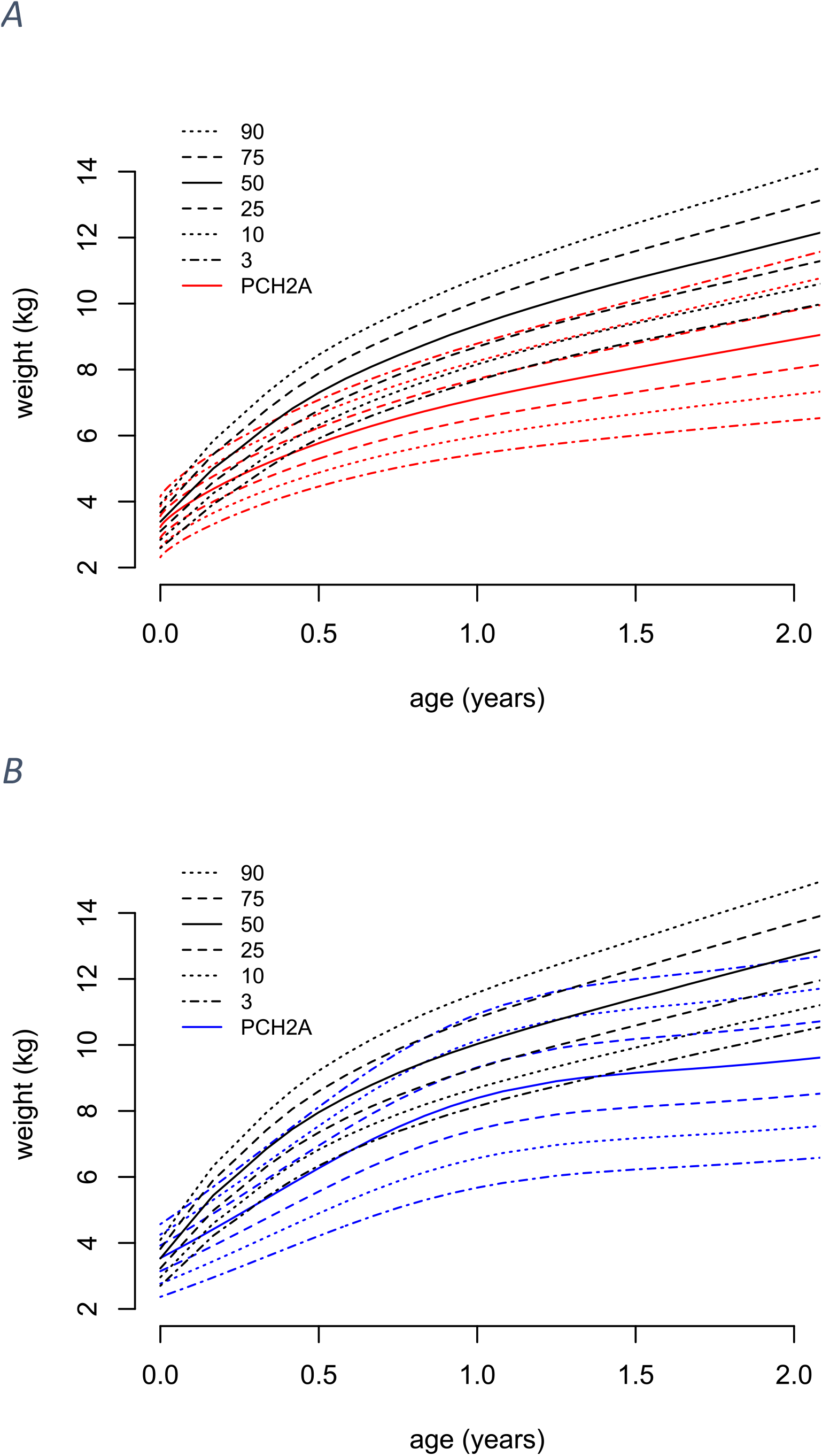
Sex-disaggregated weight charts for patients with PCH2A versus a healthy reference population (black, KiGGs) from 0-2 years; A females (red, 246 measurements, Normal model); B males (blue, 206 measurements, BCCGo model)

### BMI

Regarding BMI, reference data were available from age four months to 18 years. Overall, 440 individual measurements for BMI were collected from age 0-18 years, i.e. 248 values of female patients, and 192 of male patients. The mean number of available measurements per patient was 7 (1-18). From 4 months to 2 years, 224 measurements were available, from 2-18 years of age additional 216 measurements were available. Between 15 and 18 years of age, 11 values of female patients, and 6 measurements of male patients were included. Graphical comparison to the reference group was performed from 4 months to 2 years, statistical comparison up to the age of 5 years.

For patients with PCH2A from 4 months to 18 years of age, BMI percentiles were calculated without sex-disaggregation, but the BMI chart includes differently coloured individual measurements of female and male patients, respectively (figure 5). For patients with PCH2A age 4 months to 2 years, we provide sex-disaggregated percentiles of patients in comparison to the reference group (figure 6). At 4 months of age, mean bmi was 14.1 kg/m^2^ (SD: 1.1 kg/m^2^) in female patients, and 14.3 kg/m^2^ (SD: 2 kg/m^2^) in male patients. BMI of patients with PCH2A shows early deviation from the reference data, with statistical significance already at the first data point, i.e. at 4 months of age, in both, female and male patients with PCH2A (female: t(12)=-5.47, p=0.00014; male: t(9)=-3.14, p=0.012). BMI values remained below the reference values with statistical significance from 24 months onwards in female patients (Z=-1.97, p=0.048), and from 6 months onwards in male patients (t(17)=-4.93, p=0.00013).

**figure 5:**
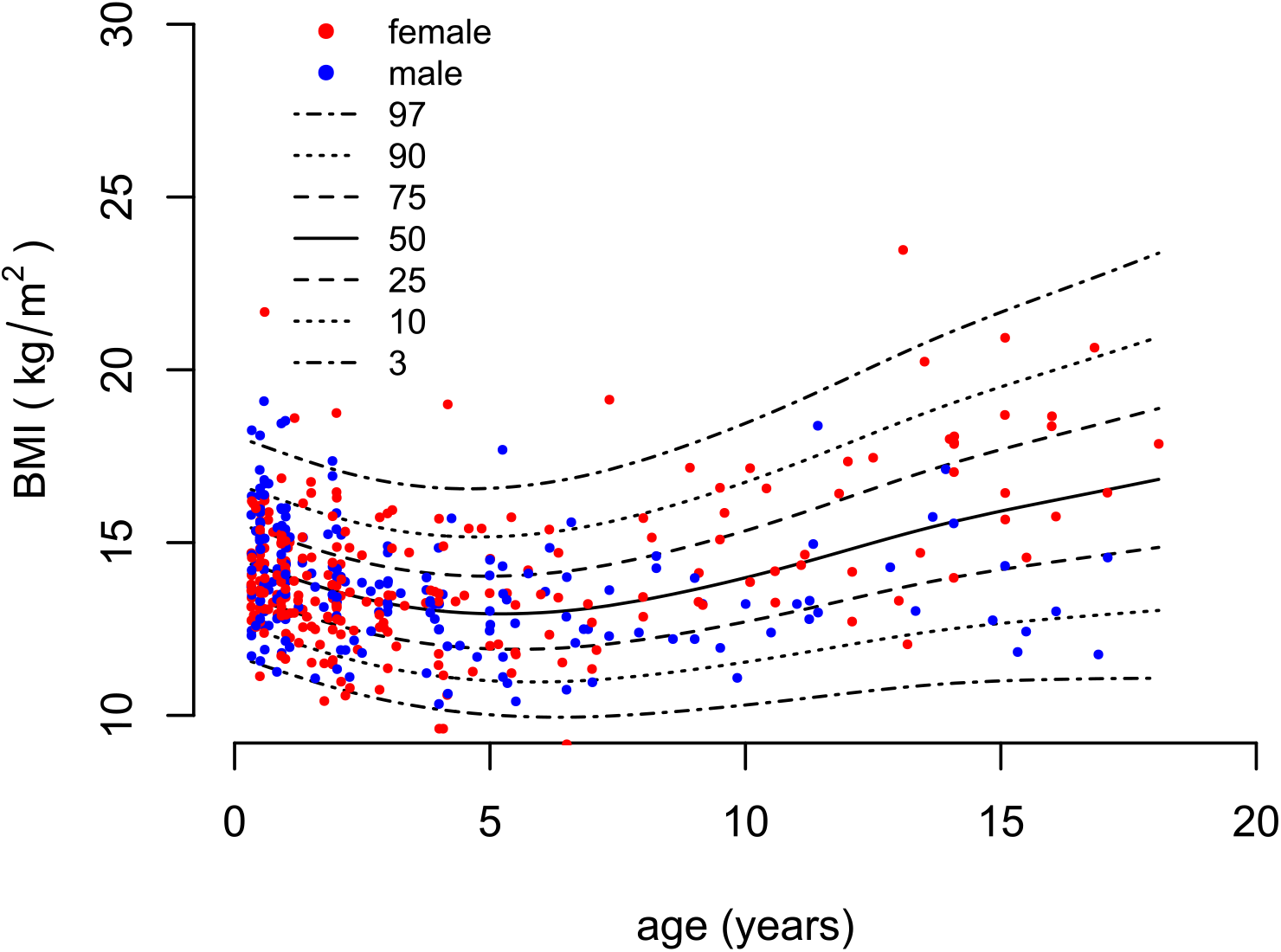
PCH2A-specific BMI charts age 4 months to 18 years. BCTo model.

**figure 6:**
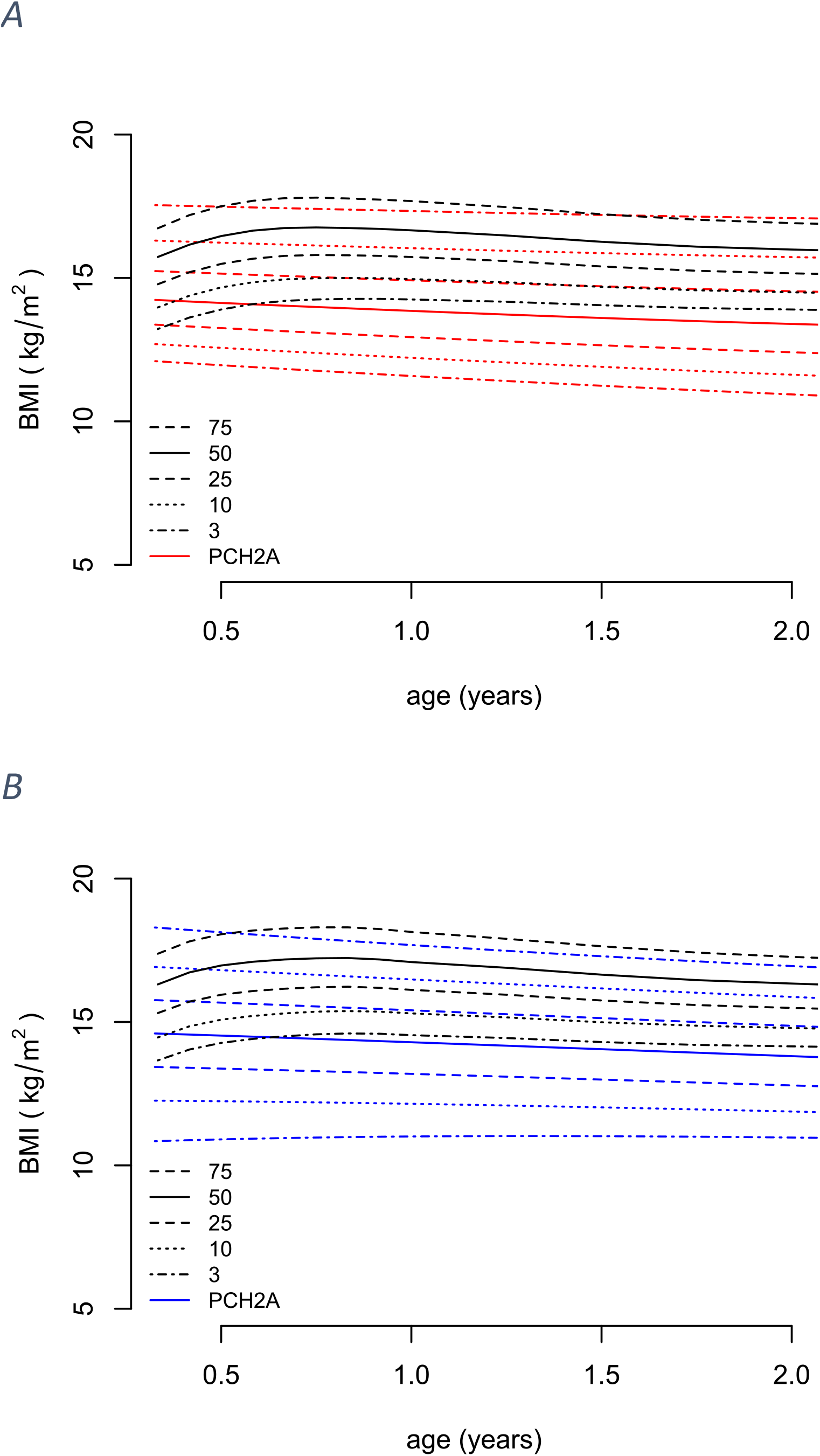
Sex-disaggregated growth charts for patients with PCH2A versus a healthy reference population (black, KiGGs) from 4 months to 2 years; A females (red, 128 measurements, BCCGo model); B males (blue, 96 measurements, BCTo model)

### Head circumference

In our cohort, 600 individual measurements for hc were collected from age 0-18 years, i.e. 320 values of female patients, and 280 of male patients. The mean number of available measurements per patient was 9.5 (1-22). From 0-2 years, 420 measurements were available, from 2-18 years of age, 180 additional measurements were available. Between 15 and 18 years of age, 8 measurements of female patients, and 6 measurements of male patients were included.

Percentiles of hc from patients with PCH2A 0-18 years of age were calculated. The chart includes differently coloured individual measurements of female and male patients with PCH2, respectively (figure 7). A graphical and statistical comparison of sex-disaggregated data with the reference group was only performed for the age group 0-2 years (figure 8). At birth, mean hc of the reference group was 34.7 cm in female and in 35.4 male patients, whereas mean hc of female and male patients with PCH2A was 33.8 cm (SD: 1.2 cm) and 34.1 cm (SD: 1.1 cm), respectively. Both values already differ significantly from the reference group at birth (female: t(31)=-4.53, p=0.000083; male: Z=-4.37; p=0.000012), and remain below reference values with increasing age. Rapid hc growth is evident during the initial years of life, and hc increases across all percentiles throughout childhood. Male individuals with PCH2A exhibit a higher hc and a higher variability of hc than female patients at the same age.

**figure 7:**
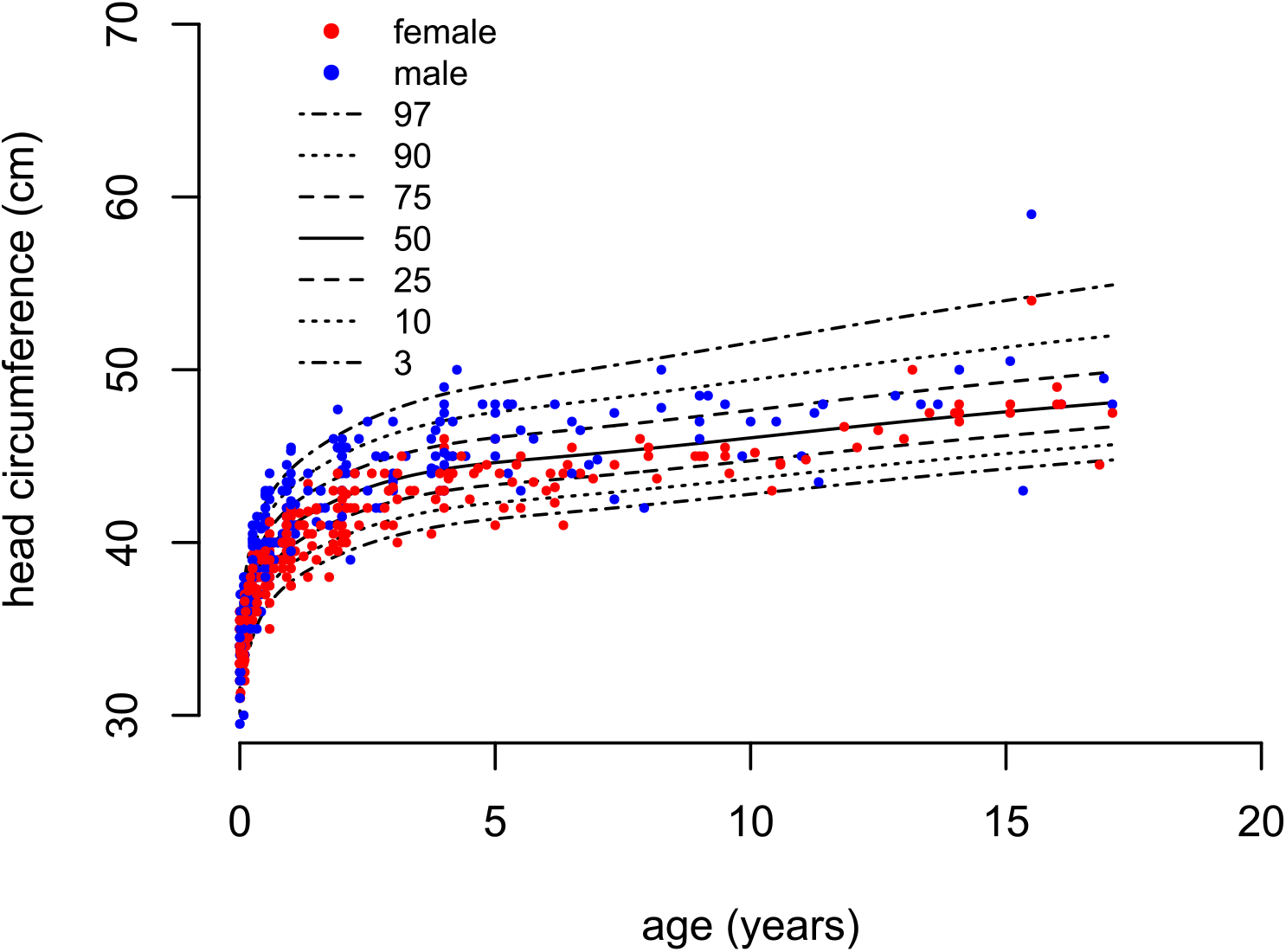
PCH2A-specific charts of head circumference (age 0-18 years). BCCGo model.

**figure 8:**
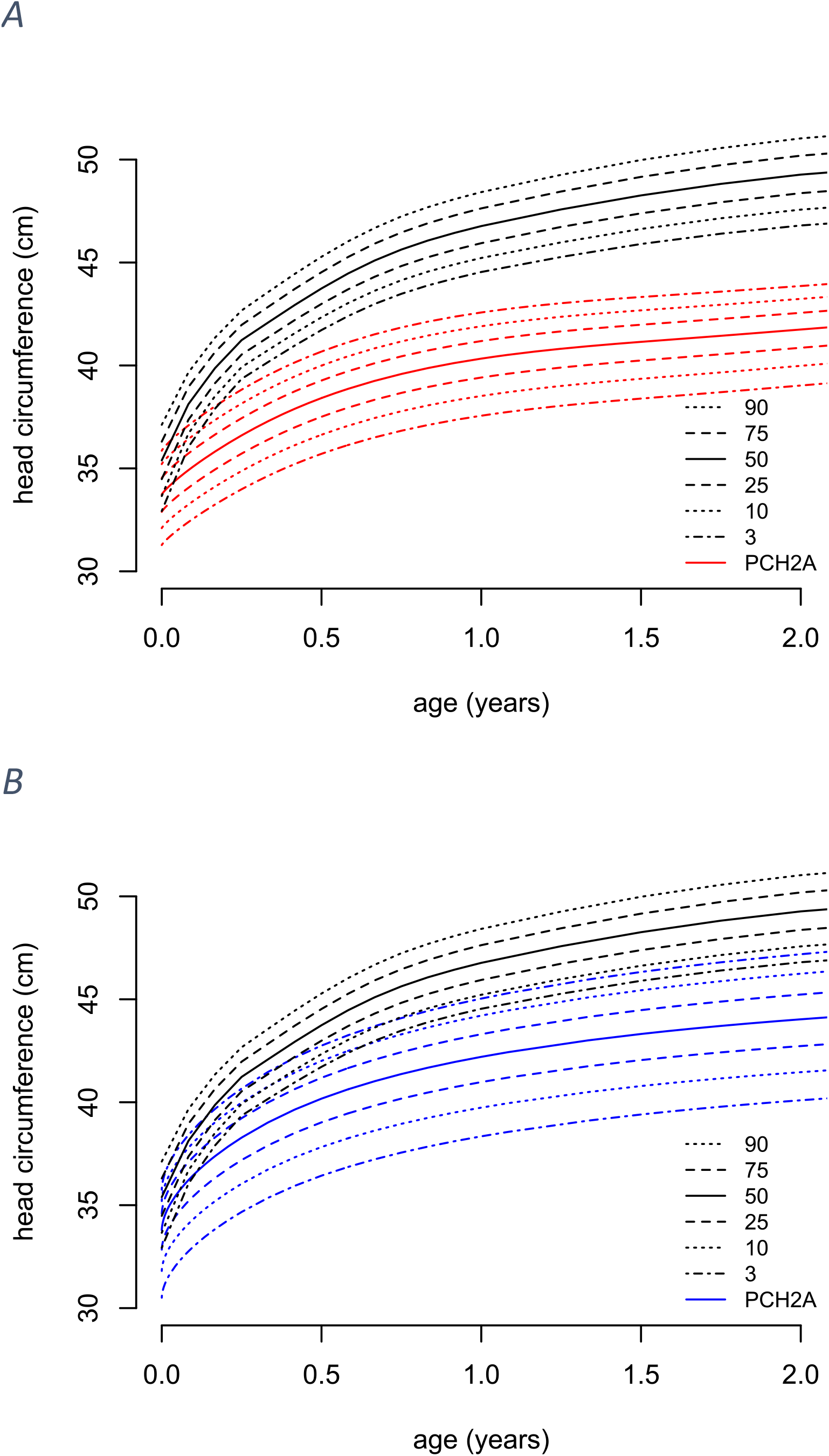
Sex-disaggregated growth charts for patients with PCH2A vs. a healthy reference population (black, KiGGs) from 0-2 years; A females (red, 224 measurements, BCTo model (k=2)); B males (blue, 196 measurements, BCCGo model)

### Feeding via percutaneous endoscopic gastrostomy tube (PEG)

Of all 65 patients with PCH2A included in the study, 37 (56, 9%) patients were provided with a PEG. Of these, 27 patients were fed at least partly via the PEG, two children were not fed via the PEG, and no data with respect to precise feeding modalities was available for the remaining eight patients with PEG. Of the 37 patients with PEG, 18 children had received at least partly feeding via PEG beginning before the age of 5 years for several years. Of these, 12/18 (66.7 %) were female, and 6/18 (33.3 %) were male. Typically, individual weight gain of patients with PCH2A showed no dramatic increase after PEG placement (Supplementary figure 1).

## Discussion

Our study aimed to provide a set of PCH2A-specific growth charts for height, weight, BMI and hc, in order to support clinical monitoring and care of affected patients. Disease-specific percentiles provide a helpful tool for patients, their families/ caretakers and treating physicians, and have already been published for several rare neurodevelopmental diseases with a similar methodological approach (13, 28, 29). Muschol et al. included 186 patients with MPS III and were the first to describe growth velocity in MPS III (13). Baer et al. calculated growth charts separately for Cockayne Syndrome (CS) 1 (49 patients) and CS2 (39 patients) as an important tool to manage nutrition in children with CS (28). Cipolli et al. provided growth charts for Shwachman-Diamond syndrome using data from 106 patients (29). To calculate PCH2A-specific growth charts, we included retrospective data of a predominantly German cohort of 65 patients with genetically proven PCH2A age 0-18 years, and applied data of a German reference population (KiGGS) as basis for comparison (14, 15).

PCH2A is a rare neurodevelopmental disorder accompanied by severe gastrointestinal symptoms, dysphagia and feeding problems (30, 31). Patients with PCH2A often fail to thrive, despite PEG-placement in many of the affected patients (5). In accordance with published data, height and weight of female and male patients with PCH2A, respectively, were still in the normal range at birth, but fell behind with increasing age (5, 32). In a previous natural history study, mean height was found to be in the normal range until the age of four years, while height of several individual children with PCH2A was already below the normal range from one year of age onwards (5). This is in line with our data, where the 50th percentile for height in both, female and male patients with PCH2A, remained above the 3rd percentile for height in the healthy reference population until the age of 2 years. In patients with PCH2A, we observed growth in height from birth to age 15 years, and progressive weight gain from birth to age 18 years. Within the first years of life, weight gain of patients with PCH2A was more affected than height. This could be at least partially explained by the severe feeding problems and gastroesophageal reflux, which are often observed in patients with PCH2A at an early age (5). BMI of patients with PCH2A decreased from birth to the age of 5 years, and then increased again. This could reflect the observation that gastrointestinal symptoms of patients with PCH2A tend to improve with increasing age (unpublished data). Moreover, more severely affected patients with PCH2A, i.e. showing more severe gastrointestinal and feeding problems at an early age, might have a higher mortality in the first years of life.

One could argue, that the presented growth charts are biased by the fact that they do not differentiate between patients receiving feeding support by PEG and those who did not. But numbers then would have been too small for meaningful analyses. In our cohort of patients with PCH2A, nearly half of all patients with PCH2A received placement of PEG during the course of the disease. It can be assumed that a PEG placement corresponds to a need in the care of the patient. Thus, those without PEG presumably still can cope with oral feeding. We chose, instead, to illustrate single patient’s weight curves before and under PEG supported feeding. It seems of note, that PEG placement showed only limited effect on weight gain in most patients. Of the patients with long-term feeding via PEG, two thirds were female. Overall, female individuals showed a better weight gain and higher BMI than male individuals, whereas hc, which is less dependent on calory intake, was higher in male patients (33). The apparently higher weight gain of female patients in comparison with male patients with PCH2A could be due to a population bias. Overall survival showed no difference between female and male patients with PCH2A.

As expected from previous descriptions, patients with PCH2 showed severe and progressive microcephaly (2, 5). Mean hc of patients with PCH2A was already below average at birth, although individual measurements of hc were often still within the lower normal range, as has already been reported (2, 5, 32). Head growth is known to be dependent on the growth of brain structures, and progressive microcephaly reflects reduced growth rate of supratentorial structures, and probably some degree of neurodegeneration over time (3, 34).

### Limitations

Discussing our data set, several limitations have to be considered. Due to the extreme rareness of the disease, the study population was relatively small, but within the range of other studies of patients with rare diseases (13, 28, 29). For individual study participants, a varying number of measurements were available. Therefore, individual growth courses affect the percentiles to a different extent. Moreover, the study included several affected pairs of siblings. For calculation of PCH2A-specific percentiles, growth data were collected from a predominantly German study cohort and compared to a German reference collective. Still, as compared to available growth percentiles of healthy children, the presented PCH2A-specific percentiles can provide an approximated reference point for other patients with PCH2A.

In our study cohort, most data were collected of patients within the first two years of life. Therefore, graphical and statistical comparison with a healthy reference group was restricted to this early age. In the older age group, there were not enough data to provide sex-disaggregated percentiles or to compare different nutritional and feeding conditions. Beyond 15 years of age, only few data could be included into our study. Only 14 patients (21.5 %) of our cohort had reached this age at the time of data acquisition, and 13 patients (20 %) had died before age 15. Especially in the older age group, PCH2A-specific growth charts should be supplemented in the future with a more solid data basis. The PCH2A-specific growth charts will nevertheless help to improve the disease management of affected patients.

## Conclusion

In our study, we provide percentiles for height, weight, BMI, and hc of patients with PCH2A as a basis for comparison of other affected patients. Despite limitations of the data, due to the extreme rarity of the disease, disease-specific growth charts will help to support and improve monitoring and care of patients with PCH2A, especially during the first two years of life, when feeding and gastrointestinal problems are often severe.

## Data Availability

All data produced in the present study are available upon reasonable request to the authors

## Literature Cited

1. Barth PG. Pontocerebellar hypoplasies: an overview of a group of inherited neurodegenerative disorders with fetal onset. Brain&Development 1993; 15(6):411–22.

2. Namavar Y, Barth PG, Kasher PR, van Ruissen F, Brockmann K, Bernert G et al. Clinical, neuroradiological and genetic findings in pontocerebellar hypoplasia. Brain 2011; 134(Pt 1):143–56.

3. Ekert K, Groeschel S, Sánchez-Albisua I, Frölich S, Dieckmann A, Engel C et al. Brain morphometry in Pontocerebellar Hypoplasia type 2. Orphanet J Rare Dis 2016; 11(1):100.

4. Budde BS, Namavar Y, Barth PG, Poll-The BT, Nürnberg G, Becker C et al. tRNA splicing endonuclease mutations cause pontocerebellar hypoplasia. Nat Genet 2008; 40(9):1113–8.

5. Sánchez-Albisua I, Frölich S, Barth PG, Steinlin M, Krägeloh-Mann I. Natural course of pontocerebellar hypoplasia type 2A. Orphanet J Rare Dis 2014; 9:70.

6. Paushkin S.V., Patel M., Furia. S., Peltz S.W., Trotta CR. Identification of a Human Endonuclease Complex Reveals a Link between tRNA Splicing and Pre-mRNA 3’ End Formation. Cell 2004; 117:311–21.

7. Sullivan PB, Lambert B, Rose M, Ford-Adams M, Johnson A, Griffiths P. Prevalence and severity of feeding and nutritional problems in children with neurological impairment: Oxford Feeding Study. Dev Med Child Neurol 2000; 42(10):674–80.

8. Claßen M, Schmidt-Choudhury A. Ernährungsprobleme und Unterernährung bei schwer neurologisch beeinträchtigten Kindern und Jugendlichen. Monatsschr Kinderheilkd 2019; 167(8):675–85.

9. Myrelid A, Gustafsson J, Ollars B, Annerén G. Growth charts for Down’s syndrome from birth to 18 years of age. Arch Dis Child 2002; 87(2):97–103.

10. Gawlik A, Gawlik T, Augustyn M, Woska W, Malecka-Tendera E. Validation of growth charts for girls with Turner syndrome. Int J Clin Pract 2006; 60(2):150–5.

11. Isojima T, Yokoya S. Development of disease-specific growth charts in Turner syndrome and Noonan syndrome. Ann Pediatr Endocrinol Metab 2017; 22(4):240–6.

12. Neumeyer L, Merker A, Hagenäs L. Clinical charts for surveillance of growth and body proportion development in achondroplasia and examples of their use. Am J Med Genet A 2021; 185(2):401–12.

13. Muschol NM, Pape D, Kossow K, Ullrich K, Arash-Kaps L, Hennermann JB et al. Growth charts for patients with Sanfilippo syndrome (Mucopolysaccharidosis type III). Orphanet J Rare Dis 2019; 14(1):93.

14. Neuhauser H, Schienkiewitz A, Rosario AS, Dortschy R, Kurth B-M. Referenzperzentile für anthropometrische Maßzahlen und Blutdruck aus der Studie zur Gesundheit von Kindern und Jugendlichen in Deutschland (KiGGS) (2003-2006); Beiträge zur Gesundheitsberichterstattung des Bundes. Robert Koch-Institut; 2013. Available from: URL: https://www.rki.de/gbe.

15. Rosario AS, Kurth B-M, Stolzenberg H, Ellert U, Neuhauser H. Body mass index percentiles for children and adolescents in Germany based on a nationally representative sample (KiGGS 2003-2006). Eur J Clin Nutr 2010; 64(4):341–9.

16. R Core Team: A language and environment for statistical computing R Foundation for Statistical Computing. Vienna, Austria; 2022. Available from: URL: https://www.R-project.org/.

17. IBM SPSS Statistics. Version 29.0.0.0 (241). Armonk, NY: IBM Corp.; 2022.

18. T.J. Cole. Fitting Smoothed Centile Curves to Reference Data. Journal of the Royal Statistical Sociaty 1988; (No.3):385–418.

19. Cole TJ, Green PJ. Smoothing reference centile curves: the LMS method and penalized likelihood. Stat Med 1992; 11(10):1305–19.

20. Stasinopoulos M, Rigby B, Voudouris V, Heller G, Bastiani F de. Flexible Regression and Smoothing: The GAMLSS packages in R [Chapter 13]; 2015.

21. R.A. Rigby, D.M. Stasinopoulos. Generalized additive models for location, scale and shape. Applied Statistics 2005; (54):507–54.

22. Cole TJ. The LMS method for constructing normalized growth standards. Eur J Clin Nutr 1990; 44(1):45–60.

23. Rigby RA, Stasinopoulos DM. Using the Box-Cox t distribution in GAMLSS to model skewness and kurtosis. Statistical Modelling 2006; 6(3):209–29.

24. Chambers JM. Graphical methods for data analysis. First edition. Boca Raton, FL: Chapman and Hall/CRC an imprint of Taylor and Francis; 2018.

25. Chambers JM, Hastie TJ. Statistical Models in S. First edition. Routledge; 2017.

26. Pan H, Cole TJ. A comparison of goodness of fit tests for age-related reference ranges. Stat Med 2004; 23(11):1749–65.

27. Stasinopoulos M, Rigby B, Voudouris V, Heller G, Bastiani F de. Flexible Regression and Smoothing: The GAMLSS packages in R [Chapter 2]; 2015.

28. Sarah Baer, Nicolas Tuzin, Peter B. Kang, Shehla Mohammed, Masaya Kubota, Yvette van Ierland et al. Growth charts in Cockayne syndrome type 1 and type 2. Eur J Med Genet 2020.

29. Cipolli M, Tridello G, Micheletto A, Perobelli S, Pintani E, Cesaro S et al. Normative growth charts for Shwachman-Diamond syndrome from Italian cohort of 0-8 years old. BMJ Open 2019; 9(1):e022617.

30. Janzarik W, Kaegeloh-Mann I, Langer T, van Buiren M, Schaefer HE, Gerner P. Spasmodic abdominal pain and other gastrointestinal symptoms in Pontocerebellar hypoplasia type 2.

31. Nguengang Wakap S, Lambert DM, Olry A, Rodwell C, Gueydan C, Lanneau V et al. Estimating cumulative point prevalence of rare diseases: analysis of the Orphanet database. European Journal of Human Genetics 2020; 28(2):165–73.

32. Steinlin M, Klein A, Haas-Lude K, Zafeiriou D, Strozzi S, Müller T et al. Pontocerebellar hypoplasia type 2: Variability in clinical and imaging findings. European Journal of Paediatric Neurology 2007; 11(3):146–52.

33. Jeong SJ. Nutritional approach to failure to thrive. Korean J Pediatr 2011; 54(7):277–81.

34. Bartholomeusz HH, Courchesne E, Karns CM. Relationship between head circumference and brain volume in healthy normal toddlers, children, and adults. Neuropediatrics 2002; 33(5):239–41.

