## supplementary figure 1 for "Growth charts for pontocerebellar hypoplasia type 2A"

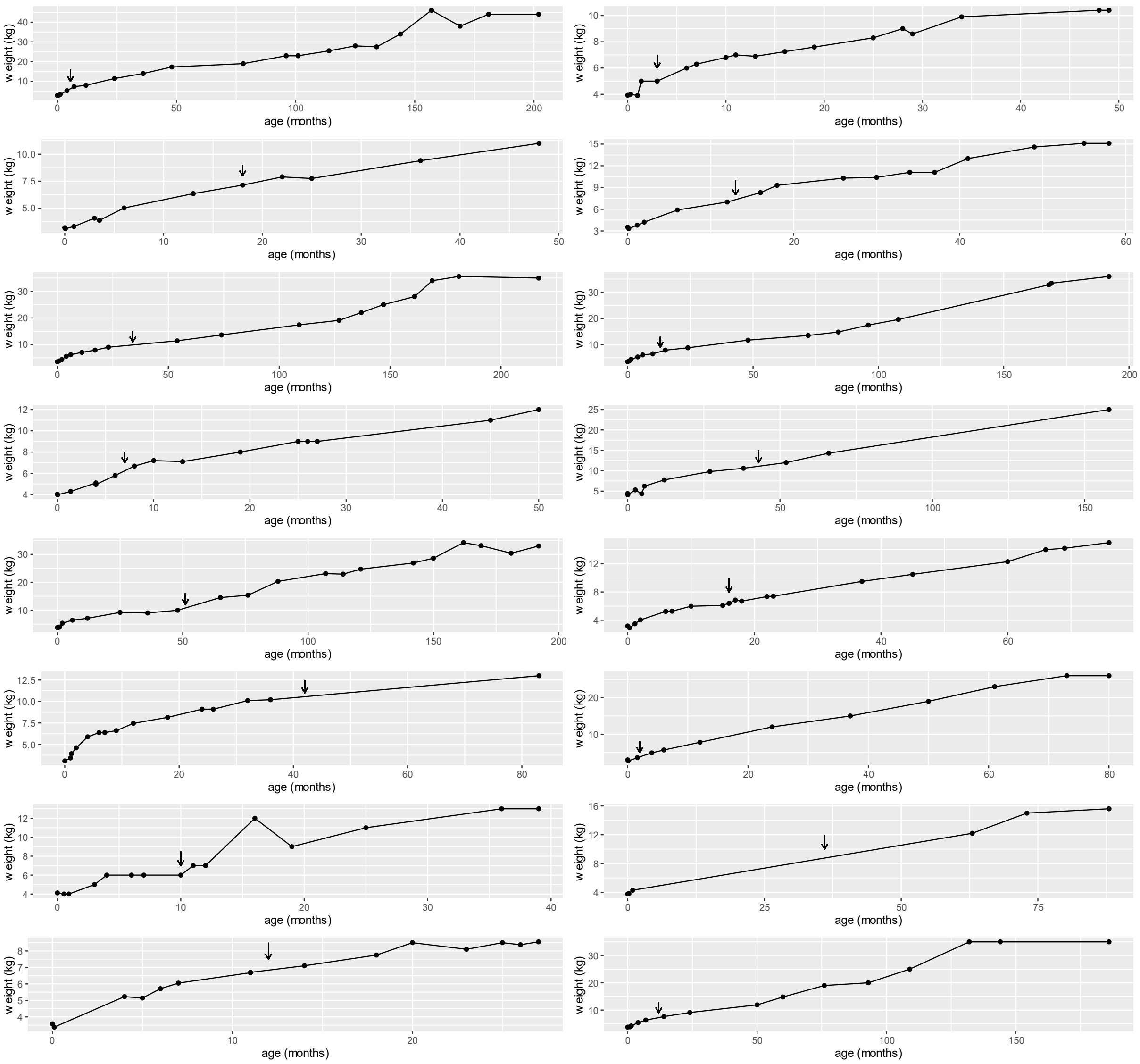


*Supplementary figure 1: Individual weight trajectories before and after PEG placement (indicated by arrows) for children who received PEG placement before the age of 5 years with long-term feeding via PEG.*
